# Sleep disturbance in people with anxiety or depressive disorders over 30 years, and the influence of personality disorder

**DOI:** 10.1101/2024.04.04.24304972

**Authors:** Jacob D King, Min Yang, Helen Tyrer, Peter Tyrer

## Abstract

**Objectives:** Sleep disturbance is commonly reported by people with anxiety, depressive and personality disorders, but longitudinal studies exploring the interplay of the three with disturbed sleep have not previously been described.

**Methods:** In this study sleep disturbance was examined among 89 patients initially presenting with anxiety or depressive disorders who provided follow-up at 12 and 30 year timepoints in The Nottingham Study of Neurotic Disorder. Multiple regression models were used to identify factors most predictive of poor sleep, and changes in sleep quality over time.

**Results:** There were strong associations between poor sleep and contemporaneous severity of personality disorder and the presence of other mental disorders at 12 and 30 years follow-up, but not with disorder presence at other time points. Improvements in personality disorder were associated with improvements in sleep between time points, and attenuated the positive unadjusted effects of recovery from anxiety or depressive disorders to insignificance. Relapse into further episodes of mental disorder predicted poorer sleep, whereas worsening personality disorder was not predictive of significant changes when adjusting for other factors.

**Conclusions:** This study demonstrates the complex interplay between anxiety, depressive and personality disorders and sleep disturbance over a long follow-up period. Future research might look to examine the relationship between personality disorder and disturbed sleep with interventional studies and by integrating personality trait research.

## 1.0 Introduction

Sleep disturbance is a common feature of both anxiety and depressive disorders (1, 2). There is a well established bi-directional relationship, with anxiety and depression symptoms contributing to poor quality sleep, and where subjective sleep disturbance predicts the onset of depressive (3) and anxiety disorders (4, 5, 6), and hinders recovery from both (7, 8). Yet previous studies have identified several factors which confound and mediate these observations, and personality traits have long been in consideration as predisposing to, and maintaining sleep disturbance and anxious and depressive symptoms (9). For example high trait neuroticism and low conscientiousness have been shown consistently to be associated with worse subjective sleep quality (10, 11, 12, 13), and the impact of sleep disturbance on depressive illness is tempered significantly when adjusting for these personality traits (6). In a further study among older adults correlations between these personality factors and sleep were mediated by measures of depression (14).

Similarities exist in aspects of sleep quality between many psychopathological groupings, notably in facets of pre-sleep hyperarousal (15), increased sleep onset latency, and poor sleep continuity and efficiency (3, 16). Moreover there are strong bi-directional associations between sleep disturbance and core clinical features of both personality disorder and common mental disorders, particularly emotion regulation (17, 18), self-harming behaviours (19), and drug and hazardous alcohol-use (20). Meta-analysis (21) failed to identify discriminatory sleep characteristics between mental health disorders, concluding instead that a transdiagnostic sleep disturbance dimension is common to many disorders (22). But while sleep disturbance features as a core symptom of anxious and depressive disorders in both DSM and ICD diagnostic classifications, it is not described as a feature of personality pathology despite its prevalence and likely influence in view of its importance in other mental disorders (16, 23, 24, 25, 26). Increasing severity of personality disorder appears to be correlated with the severity of subjective sleep disturbance (27), and poor sleep is associated with patients not recovering from core symptoms of borderline personality disorder (BPD) (28). Review studies exploring the sleep profiles of people with BPD diagnoses highlight the importance of sleep context, that sleeping alone might activate feelings of social isolation, abandonment, and rejection, and which may in part explain why there appears to be differences in observed sleep quality between the measured sleep setting (29).

The Nottingham Study of Neurotic Disorder (NSND) began recruitment in 1983 as a three-arm randomised controlled trial of 6 weeks of cognitive behavioural therapy, medication (either dothiepin, diazepam or placebo), or self-help interventions for 210 people presenting to primary care mental health services having been diagnosed with either generalized anxiety disorder, panic disorder or dysthymic disorder using a standard interview (30), or a combination of these (31). Alongside longitudinal assessments of mental state disorders the study aimed to explore the implications of co-occurring personality pathology on long-term trajectory and treatment outcomes. To date the NSND has demonstrated the dynamic nature of personality disorder severity across a 30 year life course (32, 33). Of particular importance was the finding that personality disorder appears to worsen or resolve for some individuals over time. Sleep quality, which is known to also change progressively over one’s lifetime (34) has yet to be examined for its contribution to this effect. While a handful of cross-sectional approaches have been taken to explore the relationship between sleep disturbance, anxiety, depression and personality disorder (23, 28), we are unaware of any longitudinal assessments.

Poor sleep quality is common among people diagnosed with personality disorder (35), and there appears to be short and long-term associations with suicide ideation and attempt (36, 37, 38). It has been suggested that poor sleep quality represents a “warning sign” (39) of crisis period, and could be an indicator of psychopathological complexity. Therefore it was deemed clinically important to consider how poor sleep quality as a symptom relates to, and moves in step with mental health problems over time. We therefore aimed to test the hypotheses that:

1. Sleep disturbance scores vary over a 30-year follow-up period, and will be negatively predicted by age and the presence of mental disorder.
2. The severity level of personality disorder will correlate with poor sleep at each time point.
3. The presence of personality disorder or the General Neurotic Syndrome (GNS) will attenuate the improvements in sleep associated with recovery from anxiety or depressive disorder over time.
4. Trial interventions with medication, cognitive behavioural therapy or self-help administered at baseline, will not result in lasting improvements in sleep disturbance over time.

## 2.0 Materials and Methods

This study is a secondary analysis of data collected from the Nottingham Study of Neurotic Disorder between 1982 and 2019. 210 participants with anxiety or depressive disorders were recruited from primary care mental health services in Nottingham, United Kingdom, and were allocated to a parallel arm of an initial trial of 6 weeks of either diazepam (n=28), dothiepin (n=28), placebo (n=28), Cognitive-Behavioural Therapy (n=84), or a self-help programme (n=42) by constrained randomisation. Full procedural details (31), and results of the initial trial have been reported previously (33, 40). Ethical approval to follow-up participants was granted by Northampton Research Ethics Committee (12/EM/0331).

### 2.1 Measures

The primary outcome of interest was sleep disturbance, as measured by item 4 of the observer rated Montgomery-Asberg Depression Rating Scale (MADRS) at 12 and 30 years, and change in this sleep score between time points (41). Before the 12 year time-point individual item break-down of the MADRS is not available. This item requires the clinician to assess on a 7-point Likert scale “the experience of reduced duration or depth of sleep compared to the subject’s own normal pattern when well”. Scores ranging from “sleeps as usual” scoring 0, through “slight difficulty dropping off to sleep or reduced, light or fitful sleep” scoring 2, “sleep reduced or broken by at least two hours” scoring 4, and sleeping for “less than two or three hours” per night scoring maximum 6.

Co-occurring mental health diagnoses were made at 12 and 30 years follow-up by Structured Clinical Interview for DSM (SCID) by a trained clinician; minor diagnoses like adjustment disorders were excluded as per a previously published approach (33).

Specific assessments of anxiety and depressive symptoms were made at baseline, and years 2, 12 and 30 of follow-up with the Hospital Anxiety and Depression Rating Scale’s anxiety (HADS-A) and depression (HADS-D) subscales respectively (42) – these scales are self-complete measures of 7 items apiece and which have used extensively in mental health research over 40 years, and which do not enquire into sleep qualities.

The presence and severity of personality disorder was assessed by trained psychiatrists at baseline, 2, 12 and 30 years with the Personality Assessment Schedule (PAS) (43). The PAS usually takes between 45-60 minutes to complete, and assesses 24 personality attributes on an 8-point Likert scale, and is validated to give an indication of the presence or absence of PD, and a severity level (no personality disorder, the sub-clinical ‘personality difficulty’, and simple, or complex personality disorder), alongside type of personality disturbance, which is analogous to, and concordant with, contemporary ICD-11 criteria (44). The PAS maintained good interrater agreement on example case vignettes: κ=0.65 (45).

At baseline we also assessed for the presence of the General Neurotic Syndrome (46), the combination of anxiety and depressive symptoms, ‘cluster C’ personality disturbance and parental history of mixed anxious and depressive symptoms, as delineated by a score of ≥4 on the General Neurotic Scale, to explore whether the presence of this common combination of affective and personality problems had an impact on sleep after long period of follow-up.

Finally, at the 12 and 30 year follow-up periods a clinician-rated ‘recovery score’ measuring current severity of anxiety and depressive symptoms was assessed, constructed on a 4 point scale – where 4 represents full recovery from their symptoms, and 1 represents a current severe level of symptoms. A score of 4, fully-recovered, was only given if there were no significant symptoms of anxiety or depressive disorders at the time of follow-up, nor contact received for mental health concerns from the person’s primary care physician or psychiatrist since the previous time-point. Further details of all measures used in this study are reported in detail in other analyses of this cohort (33, 40).

### 2.2 Statistical analysis

These analyses are based on 89 cases from the original cohort of 210, who had complete data from baseline to 30 years of follow-up (42.8%). By this time point 71 people (33.8%) had died, and a further 50 (23.8%) were not assessed most commonly because they were no longer contactable. Attrition analyses of baseline clinical and demographic factors identified only greater age as predictive of loss to follow-up at 12 and 30 year time points. Full reporting of cohort attrition is reported elsewhere (33).

Simple correlation analyses with Spearman’s *rho* were used to identify the relationships between demographic details, personality disorder severity, diagnoses of mental health problem by DSM-III criteria, recovery from baseline anxiety or depressive disorders and sleep disturbance at each time point. Weak correlation was taken as *r*<0.4, moderate <0.7 and strong correlation ≥0.7 (47).

Sleep disturbance at time point 12, and 30 years, as well the change (sleep score at 30 years subtract score at 12 years) in sleep disturbance over the 18 year period were the 3 dependent variables used in multivariate regression models. For each dependent variable four models were conducted. These factors were entered into models in a nested approach to show possible adjustment effects of one factor on another. Model 1 examined the effects of age only. Model 2 added effects of PD severity at both earlier and current time points on current sleep disturbance. Model 3 added presence of DSM-III diagnosis at the current time, and Model 4 added a measure of recovery from baseline anxiety or depressive illness at 30 years. For sleep change over time, the changes in PD severity and DSM-III diagnosis over the same period were entered as predictors. The independent predictive effects of personality status, DSM-III status and long-term recovery from the anxiety or depressive disorders and their association with disturbed sleep were hence estimated. In the same vein we tested how changes in PD severity and the emergence/loss of DSM-III diagnosis were associated to changes in sleep disturbance during the same period. Regression analyses were also used to test effects of initial trial allocation in treatment group on sleep disturbance at later times. IBM SPSS v19 was used to perform all analyses.

## 3.0 Results

Female and male distribution in this cohort was 70.8% and 29.2%; there was a mean age of 30.8 ± 8.6 years at baseline. Sleep disturbance was common, with only 51.7% reporting sleeping as usual (scoring 0 on MADRS item 4) at the 12 year time-point, and a minority 43.2% at 30 years. Table 1 displays descriptive statistics of key variables for the study. At 30 years, 42.7% of the cohort had fully recovered, indicating that they no longer met DSM-III diagnostic criteria for a mental disorder or those for a personality disorder under assessment by the PAS.

**Table 1.** Descriptive statistics of key variables used in the study.

| Variables |  | Baseline | 2 years | 12 years | 30 years |
| --- | --- | --- | --- | --- | --- |
| Age | Mean ± SD | 30.8 ± 8.6 | | | |
| DSM | Presence | 89 (100.0) |  | 43 (48.3) | 45 (50.6) |
|  | Absence |  |  | 46 (51.7) | 44 (49.4) |
| DSM change<br>(12-30 years)<sup>a</sup> | No change |  |  | 55 (61.8) |  |
|  | Improved |  |  | 16 (18.0) |  |
|  | Worsened |  |  | 18 (20.2) |  |
| PD<sup>b</sup> | None | 35 (39.3) | 33 (37.1) | 38 (42.7) | 30 (33.7) |
|  | Difficulty | 16 (18.0) | 16 (18.0) | 16 (18.0) | 16 (18.0) |
|  | Simple | 27 (30.3) | 13 (14.6) | 19 (21.3) | 19 (21.3) |
|  | Complex | 9 (10.1) | 8 (9.0) | 16 (18.0) | 24 (27.0) |
|  | Missing | 2 (2.2) | 19 (21.3) |  |  |
| PD change<br>(12-30 years)<sup>c</sup> | No change |  |  | 63 (70.8) |  |
|  | Improved |  |  | 9 (10.1) |  |
|  | Worsened |  |  | 17 (19.1) |  |
| Initial group | Dothiepin | 10 (11.2) |  |  |  |
|  | CBT | 40 (44.9) |  |  |  |
|  | Self-help | 17 (19.1) |  |  |  |
|  | Other drug | 22 (24.7) |  |  |  |
| Fully recovered | Yes |  |  |  | 38 (42.7) |
|  | No |  |  |  | 51 (57.3) |
| GNS≥6 | Yes | 28 (31.5) | | | |
|  | No | 60 (67.4) |  |  |  |
|  | Missing | 1 (1.1) |  |  |  |
| Sleep quality score | Mean ± SD | | | 1.47 ± 1.78 | 1.47 ± 1.62 |

DSM – Diagnosis under the Diagnostic and Statistical Manual of Mental Disorders IIIrd edition. PD – Personality Disorder. GNS – General Neurotic Syndrome.
<sup>a</sup>No change in DSM diagnosis is defined for those with either DSM absence or DSM presence at both 12 and 30 years; Improved DSM describes those who change from DSM presence at 12 years to DSM absence at 30 years; Worsened DSM is for those with DSM absence at 12 years changing to DSM presence at 30 years.
<sup>c</sup>PD change is derived from the two PD groups at 12 and 30 years, using the same approach as for the DSM changes.

Correlation analysis reported in Table 2 demonstrates a number of findings. Firstly, that sleep disturbance score at 12 years was moderately positively correlated with sleep disturbance 18 years later. Second, that the age of patients had no correlation with sleep disturbance at any time point. Third, while PD severity level measured at baseline and the 2 year time point was not associated with disturbed sleep at any later time point, contemporary PD status had a strong positive association with sleep disturbance at the time when both were assessed. Fourth, the presence of a DSM diagnosis at 12 years was only very weakly associated with also meeting criteria for a DSM-III diagnosis at 30 years (*r*=0.24), but did have a strong and positive correlation with the time-matched PD severity level, and sleep disturbance. Recovery from anxiety and depression between 12 and 30 years follow-up was significantly related to improved sleep and to improved PD status. Higher scores on the general neurotic syndrome scale at baseline were only weakly correlated with sleep disturbance at 12 years and became insignificant at 30 years. Participants’ GNS baseline score had little correlation with PD severity level at all times, nor with DSM diagnoses, and was therefore not considered for inclusion in further analyses.

**Table 2.**
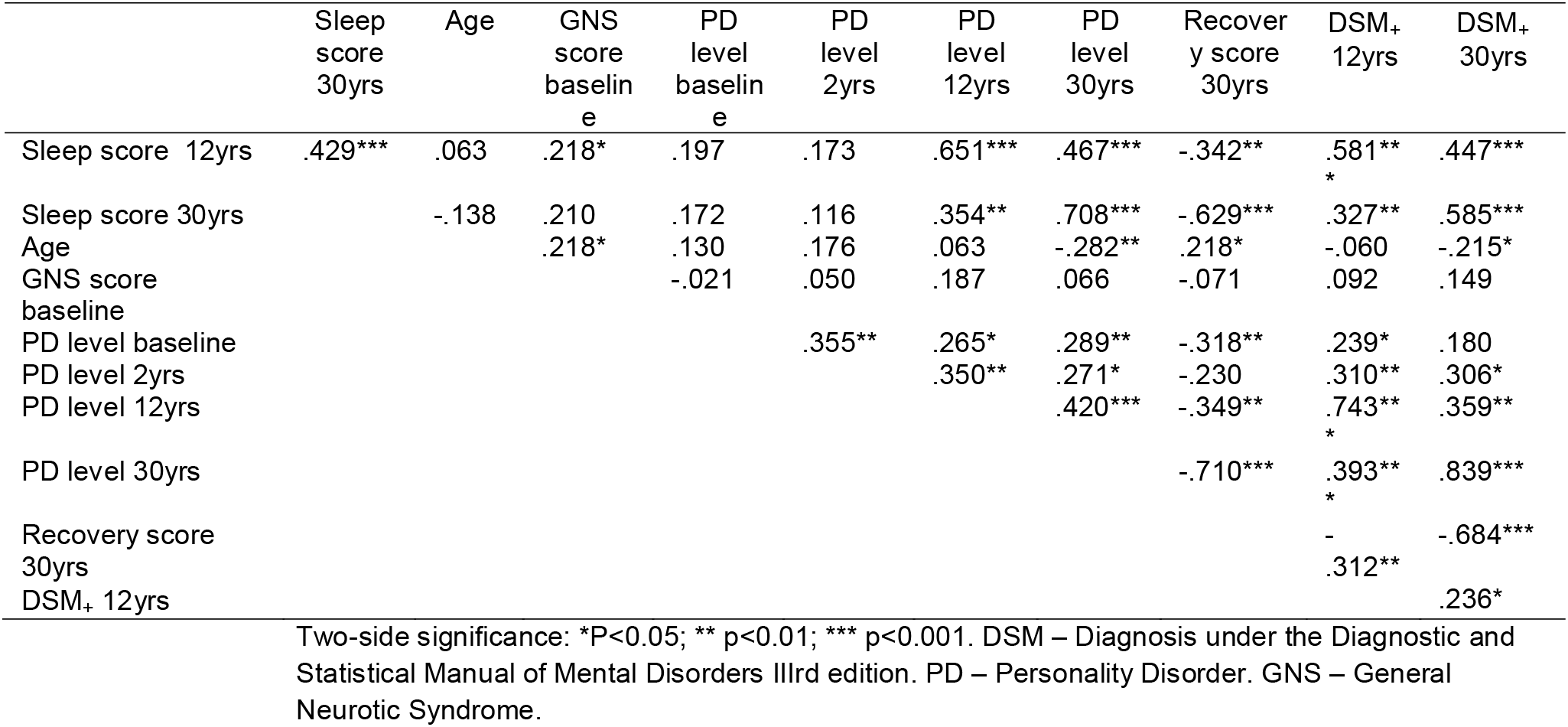
Simple correlations among sleep quality score and age, GNS score, PD severity level as well as recovery score of anxiety-depression (Spearman’s coefficient rho)

### 3.1 Anxiety, depression and sleep disturbance

Simple correlation analysis between anxiety and depression and sleep disturbance at 12 and 30 years demonstrated increasing strengths of associations with contemporaneousness: there were increasingly strong associations between anxiety and/or depressive scores and poor sleep at the times when both were measured. Additionally, a long-term predictive effect of early anxiety or depression score to later disturbed sleep emerged, so while anxiety and depression scores at 2 years’ follow-up moderately predicted sleep disturbance at 12 years’ follow-up, it had a strong predictive effect from 12 years to 30 years. The negative relationship between sleep disturbance and recovery level over time showed a mirror image of the effect of anxiety and depression score on sleep disturbance. Given that measures of anxiety and depression were strongly correlated at all time points, to maintain the independence of covariates and to simplify the analysis only the recovery variable was used in later multivariate regression analysis as predictors of sleep disturbance.

### 3.2 Effects of initial treatment allocation

Table 3 outlines the 12 and 30 year mean sleep scores of patients by their initial trial allocation arm. There was no sign of benefit from any of the initial allocated interventions on differential sleep disturbance at long-term follow-up, and patients with PD or DSM presence at both 12 and 30 years had significantly worse sleep regardless of early treatment group.

**Table 3.** Sleep quality scores by initial treatment allocation and by long follow-up time points.

|  | Drugs |  | CBT |  | Self-help |  | P value<sup>a</sup> |  |
| --- | --- | --- | --- | --- | --- | --- | --- | --- |
|  | PD. | PD<sub>+</sub> | PD. | PD<sub>+</sub> | PD. | PD<sub>+</sub> | PD or DSM | Treatments |
| **12 years** |  |  |  |  |  |  |  |  |
| N | 24 | 8 | 34 | 6 | 15 | 2 |  |  |
| mean±SD | 1.17±1.61 | 3.88±1.25 | 1.24±1.76 | 1.83±1.84 | 0.93±1.49 | 2.50±0.71 | **0.000** | 0.637 |
| **30 years** |  |  |  |  |  |  |  |  |
| N | 26 | 6 | 25 | 14 | 13 | 4 |  |  |
| mean±SD | 0.96±1.40 | 2.50±1.38 | 0.72±0.89 | 3.36±1.50 | 1.31±1.70 | 1.75±1.71 | **0.000** | 0.994 |
|  | DSM- | DSM+ | DSM- | DSM+ | DSM- | DSM+ |  |  |
| **12 years** |  |  |  |  |  |  |  |  |
| N | 16 | 16 | 19 | 21 | 11 | 6 |  |  |
| mean±SD | 0.88±1.50 | 2.81±1.83 | 0.26±0.65 | 2.29±1.90 | 0.36±0.81 | 2.50±1.52 | **0.000** | 0.280 |
| **30 years** |  |  |  |  |  |  |  |  |
| N | 16 | 16 | 17 | 22 | 10 | 7 |  |  |
| mean±SD | 0.63±1.03 | 1.88±1.67 | 0.41±0.71 | 2.64±1.62 | 0.50±0.85 | 2.71±1.70 | **0.000** | 0.731 |

<sup>a</sup> Wald test of model effects by means of linear multivariate regression analysis.
DSM – Diagnosis under the Diagnostic and Statistical Manual of Mental Disorders IIIrd edition. PD – Personality Disorder. SD – Standard Deviation.

### 3.3 Predicting improvement in sleep

Table 4 presents the estimated impacts of age, PD severity, PD severity change, DSM-III diagnosis presence, DSM-III diagnosis change and recovery status on sleep disturbance at 12 years, at 30 years and change of sleep disturbance between 12 and 30 years respectively.

**Table 4.**
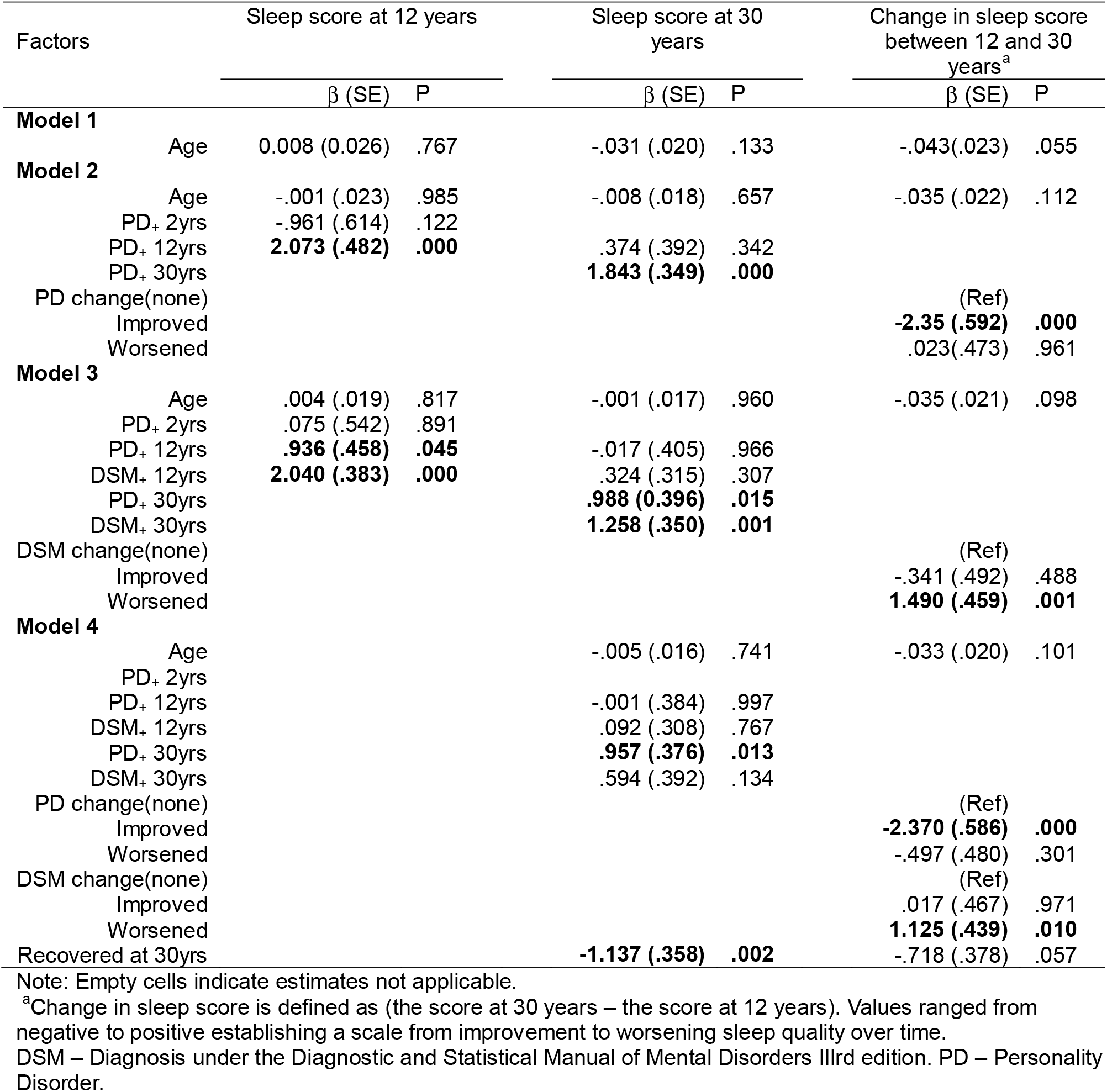
Predictive factors for improvement of sleep quality using multiple regression models.

| Factors | Sleep score at 12 years |  |  | Sleep score at 30 years |  |  | Change in sleep score between 12 and 30 years<sup>a</sup> |  |  |
| --- | --- | --- | --- | --- | --- | --- | --- | --- | --- |
| | | $\beta$ (SE) | P | | $\beta$ (SE) | P | | $\beta$ (SE) | P |
| **Model 1** |  |  |  |  |  |  |  |  |  |
| Age |  | 0.008 (0.026) | .767 |  | -.031 (.020) | .133 |  | -.043(.023) | .055 |
| **Model 2** |  |  |  |  |  |  |  |  |  |
| Age |  | -.001 (.023) | .985 |  | -.008 (.018) | .657 |  | -.035 (.022) | .112 |
| PD+ 2yrs |  | -.961 (.614) | .122 |  |  |  |  |  |  |
| PD+ 12yrs |  | **2.073 (.482)** | **.000** |  | .374 (.392) | .342 |  |  |  |
| PD+ 30yrs |  |  |  |  | **1.843 (.349)** | **.000** |  |  |  |
| PD change(none) |  |  |  |  |  |  |  | (Ref) |  |
| Improved |  |  |  |  |  |  |  | **-2.35 (.592)** | **.000** |
| Worsened |  |  |  |  |  |  |  | .023(.473) | .961 |
| **Model 3** |  |  |  |  |  |  |  |  |  |
| Age |  | .004 (.019) | .817 |  | -.001 (.017) | .960 |  | -.035 (.021) | .098 |
| PD+ 2yrs |  | .075 (.542) | .891 |  |  |  |  |  |  |
| PD+ 12yrs |  | **.936 (.458)** | **.045** |  | -.017 (.405) | .966 |  |  |  |
| DSM+ 12yrs |  | **2.040 (.383)** | **.000** |  | .324 (.315) | .307 |  |  |  |
| PD+ 30yrs |  |  |  |  | **.988 (0.396)** | **.015** |  |  |  |
| DSM+ 30yrs |  |  |  |  | **1.258 (.350)** | **.001** |  |  |  |
| DSM change(none) |  |  |  |  |  |  |  | (Ref) |  |
| Improved |  |  |  |  |  |  |  | -.341 (.492) | .488 |
| Worsened |  |  |  |  |  |  |  | **1.490 (.459)** | **.001** |
| **Model 4** |  |  |  |  |  |  |  |  |  |
| Age |  |  |  |  | -.005 (.016) | .741 |  | -.033 (.020) | .101 |
| PD+ 2yrs |  |  |  |  |  |  |  |  |  |
| PD+ 12yrs |  |  |  |  | -.001 (.384) | .997 |  |  |  |
| DSM+ 12yrs |  |  |  |  | .092 (.308) | .767 |  |  |  |
| PD+ 30yrs |  |  |  |  | **.957 (.376)** | **.013** |  |  |  |
| DSM+ 30yrs |  |  |  |  | .594 (.392) | .134 |  |  |  |
| PD change(none) |  |  |  |  |  |  |  | (Ref) |  |
| Improved |  |  |  |  |  |  |  | **-2.370 (.586)** | **.000** |
| Worsened |  |  |  |  |  |  |  | -.497 (.480) | .301 |
| DSM change(none) |  |  |  |  |  |  |  | (Ref) |  |
| Improved |  |  |  |  |  |  |  | .017 (.467) | .971 |
| Worsened |  |  |  |  |  |  |  | **1.125 (.439)** | **.010** |
| Recovered at 30yrs |  |  |  |  | **-1.137 (.358)** | **.002** |  | -.718 (.378) | .057 |

Note: Empty cells indicate estimates not applicable.
<sup>a</sup>Change in sleep score is defined as (the score at 30 years – the score at 12 years). Values ranged from negative to positive establishing a scale from improvement to worsening sleep quality over time.
DSM – Diagnosis under the Diagnostic and Statistical Manual of Mental Disorders IIIrd edition. PD – Personality Disorder.

For sleep disturbance at 12 years, meeting criteria for PD and DSM-III diagnoses at 12 years were independently associated with poor sleep: the impact of a DSM diagnosis was stronger than that of a positive diagnosis of PD. The age of patients had no impact, nor did previous PD and DSM diagnoses at the 2 year follow-up point predict sleep disturbance 10 years later. Likewise, age, PD and DSM diagnosis at 12 years did not show significant impacts on sleep disturbance at 30 years, but a PD or DSM diagnosis at 30 years was significantly and independently associated with poorer sleep at 30 years. The impact of a DSM-III diagnosis was attenuated by the effect of recovery from anxiety or depressive disorders at 30 years while they themselves predicted significant improvements in sleep disturbance. The strong negative predictive effect of PD status on sleep disturbance at 30 years was resistant to controlling for the benefit of recovery from anxiety or depression.

In relation to sleep disturbance change over 18 years, age alone had a borderline negative effect which was weakened by the strong effect of improving PD severity. The same finding was evidenced by the relationship between changes in sleep disturbance and presence of DSM-III diagnosis, showing that developing a new DSM-III diagnosis was significantly associated with worsening sleep over time. The effect of recovery on change of sleep disturbance was borderline, which did not affect the co-varied relationship between PD or DSM changes and change in sleep disturbance.

## 4.0 Discussion

This study highlights the associations of anxiety, depressive and personality disorders with sleep over a long follow-up period. The presence of sleep disturbance was high in this population. For comparison, a study using the same method of assessment of sleep disturbance in a similarly aged group of individuals from a general population sample in Sweden (mean age 67.5, s.d 7.9) reported sleep disturbance in only 19.7%, compared with 56.8% in the present cohort at the 30 year follow-up period (mean age 60.8, s.d 8.6) (48).

There were two notable and novel findings from this analysis. Firstly, because we used an approach of personality assessment analogous to ICD-11 Personality Disorder severity assessment, these results were able to demonstrate a gradient between increasing severity of personality disorder and poorer sleep duration and depth in the same individuals at different stages of their life. To date, work on this topic had used cross-sectional methods or dichotomous PD outcome measures (27, 28). Moreover, when controlling for contemporary personality disorder severity, personality disorder presence at other time points was not associated with sleep disturbance, suggesting that the degree of sleep disturbance moves in-step with severity of personality pathology across the life course. Together these observations highlight the role of poor sleep as an independent indicator of current personality disorder severity, rather than there being a persistent longitudinal effect of a historical personality disorder diagnosis. In the McClean Study of Adult Development, an American longitudinal study of 290 people with (severe Borderline) personality disorder, at the 16 year follow-up mark, patients who reported disturbed sleep were significantly less likely to have achieved recovery from their BPD symptoms (28). Authors of that study had concluded that poor sleep may therefore hamper recovery from BPD. Our study builds on this by demonstrating changes in sleep disturbance follow in-step with personality status. We might however suggest, that while both studies show that sleep disturbance may indicate contemporary personality status, the mediation pathway remains unclear: does personality decompensation lead to poor sleep as a symptom (among others) of poor personality functioning, does poor sleep in part perpetuate personality decompensation, or do factors which both precipitate sleep disturbance also precipitate mental health ‘crisis’?

Secondly, these results also demonstrate that poor sleep is positively associated with severity of personality disorder independent of cooccurring anxiety or depressive illness. While unadjusted models confirm preestablished findings that recovery from mental illness is associated with improved sleep outcomes, the present results suggest that these benefits are associated instead with improvements in cooccurring personality disorder. Correspondingly, recovery from anxiety and depressive disorders were not associated with improvement in sleep disturbance as might have been expected when improvements in personality disorder were taken in to account. The recurrence, or development of new mental state disorder however was shown to be strongly associated with a worsening sleep disturbance, while worsening personality disorder status was not. These findings suggest that the previously conceptualised bi-directional relationship of anxiety and depressive disorders and sleep is more complex than currently understood, and challenge the notion that treating depressive or anxiety disorders without addressing personality disturbance will improve sleep. In this way findings mirror those which indicate personality factors as mediating the relationship between common mental disorders and sleep quality (12, 14).

There are several mechanisms by which sleep disturbance is linked with personality disorder, but none have achieved wide-spread acceptability in the literature. Some evidence has pointed to a shared aetiology from temperament, childhood adversity, maladaptive parenting, nightmares, other primary sleep disorders, hypothalamic-pituitary axis dysfunction (49), and circadian rhythm disturbance (26). Other authors highlight causative or mediating factors of PD which contribute to disturbed sleep including drug and alcohol use, emotional dysregulation (50), or cognitive distortions (51). Similarly, reverse causation is postulated given that disturbed sleep is associated with emotional lability, impulsivity and self-harming behaviours (52).

In light of this study’s findings that an improving PD status is associated with improved sleep, we propose that addressing personality disturbance with established interventions could lead to improved sleep for patients resistant to other approaches. Moreover, the reverse may also stand, and improving sleep by targeted intervention could herein have benefit to the core outcomes of those with personality disorder, as it is beginning to be shown in many mental disorders (53), as well as chronic pain (54) and fibromyalgia (55). Experimental studies which improve sleep quality with established interventions psychological and pharmaceutical, and measure personality disorder severity and associated outcomes as well as that which integrates research on personality trait profiles and cooccurring mental state disorders would be of benefit to clarify this relationship.

### 4.1 Limitations

The main limitation of this study was that the outcome measure for sleep disturbance was a single item, and while items in the MADRS are constructed on Likert scales and treated as continuous variables (56) these individual Likert-type items have not been assessed for their construct validity, or suitability for assessment of a unifying concept as a continuous variable. Moreover this item is based on observer judgement rather than objective measures, and relates only to aspects of duration and depth; these analyses are further limited insofar as individual MADRS items are not available at baseline. Additionally, the time-points available may not be the most optimal for capturing changes in sleep, mental state and personality profiles, and naturally more frequent assessments would give a sharper image of the trajectory of these variables.

It would be pertinent for future studies of sleep disturbance in personality and mental state disorders to use validated assessment scales or objective outcomes which assess multiple facets of sleep quality. In addition, given the complexity of the relationship between anxiety, depressive and personality disorders and sleep disturbance, there are several key confounders through the timeline for which we have not been able to account, particularly current medication (especially sedative and hypnotic agents), drug and alcohol use, major life events, primary sleep disorders, and physical health comorbidities.

## 5.0 Conclusions

Sleep disturbance may be a “neglected symptom” (50) of personality disorder, given that these results have shown a strong, temporal and graded relationship. Recent developments in diagnostic criteria for personality disorder better facilitate work which might now look to include personality traits, and severity of personality dysfunction into the established literature base tackling the links between common mental health disorders and sleep. Future work in this field may explore whether sleep disturbance can be used as a severity indicator of personal disorder, or indeed whether improving sleep quality improves core outcomes of personality disorder and *vice versa*.

## Author contributions

All authors meet ICJME criteria for authorship. All authors have edited and approved this article for submission. JDK and PT conceived of the hypotheses. MY conducted all statistical analyses. HT and PT collected raw data. JDK, MY and PT drafted the initial manuscript.

## Data availability

Data relating to these analyses are available from the corresponding author on reasonable request.

## Ethics statement

The original study was approved by the Nottingham Ethics Committee in 1983, and ethical approval to follow-up participants was granted by Northampton Research Ethics Committee (12/EM/0331).

